# Socioecological benefits of academic greenspace for human health, plant, and pollinator diversity: a mixed-method study protocol

**DOI:** 10.1101/2024.12.30.24319778

**Authors:** T.O. Kehinde, A. Akinsulore, B.O. Mapayi, V.O. Okorie, M. Oziegbe, C.C. Ndiribe, O.A. Adedoja, I. Balogun

## Abstract

**Introduction:** Significant risks to the health of humans and ecosystems are posed by environmental pollution, urban warming, and fragmentation, which are primary urban factors that contribute to the deterioration of urban ecosystems. The purpose of this multidisciplinary study is to compare greenspaces in university campuses and the host cities in order to assess how valuable they are for promoting human and ecosystem health in Nigeria.

**Methods:** Mixed-methods research that will be conducted in five tertiary institutions and their host cities in southwestern Nigeria. Based on the objectives, the study is divided into four work packages (WP). WP 1 will use suitable sampling traps and scheduled field observations to quantify the diversity of plants and pollinators. Quantitative evaluation of well-being and mental health will be done in WP 2. In WP 3, the relationship between ecosystem health and mental health will be examined and in-depth interviews will be used to explore the socioecological perceptions and interaction of people with specific indicators of ecosystem health in greenspace. In WP 4, a nature-based intervention will be developed and evaluated in a pilot study to determine its feasibility and acceptability.

**Results:** The 2023 TETFund National Research Fund Intervention provided funding for this work, which is currently ongoing.

**Conclusion:** This project will provide scientific knowledge to support evidence-based policy for relevant stakeholders and regulatory organisations, with the goal of promoting greenspace infrastructure in Nigerian tertiary institutions.

## Introduction

Human well-being is significantly impacted by urbanisation, which also has an effect on the public, mental, and economic health of urban ecosystems. Urbanisation is a growing global problem to human and environmental health due to increased human migration into urban cities [1]. Urbanisation is closely linked to key drivers of global change, many of which are concentrated in metropolitan areas. These drivers include urban warming, environmental pollution, fragmentation, and habitat loss. These factors affect the distribution of biodiversity, human livelihood, and the socioecological relationships that sustain ecosystem health and human well-being in urban landscapes.

Urban greenspaces hold the potential to provide nature-based solutions, as noted by [2], especially in light of the expected growth in urban land cover and population size, particularly in tropical regions [3]. But more research is still needed to determine how greenspaces improve the health of urban residents by fostering a diverse population of ecologically significant species, including pollinators and plants, as well as mitigating the negative consequences of urbanisation.

University campuses are widely acknowledged to provide a chance for a potentially healthy greenspace framework that incorporates advantages for human and environmental health inside the urban matrix. Moreover, they serve as a sentinel landscape for the prompt exploration of the possible advantages of greenspace for human health and biodiversity. This is especially crucial in some developing countries, like Nigeria, where university campus landscapes are better designed to integrate greenspaces than host city landscapes, where greenspaces are primarily on the periphery and are more closely related to rural than urban environments [4].

Studies examining how pollinator health affects human health through their direct and indirect effects on plant diversity and greenspace quality are generally lacking [5]. This is despite the wealth of evidence suggesting pollinator and plant diversity as a limiting factor of ecosystem health and the quality of greenspaces [6,7,8]. This is an important factor to take into consideration when managing greenspaces since other elements of the environment (for example pests or invasive species) may not be the best indicators of how beneficial greenspace may be for human health.

Therefore, multidisciplinary research is required to determine the relationships between ecosystem health in urban greenspaces and human health using appropriate socioecological context, such as that found in university greenspaces. The aim of this study is to determine the value of urban greenspaces on university campuses versus their host cities in promoting ecosystem and human health. The specific objectives are to assess the diversity of pollinators, plants, pollinator-plant interactions and landscape proportion of green area as indicators of ecosystem health in greenspaces on university campuses and their host cities; evaluate the self-perceived mental health indices of people living close to greenspaces on university campuses and in the host cities; determine the relationship among diversity of pollinators, plants, pollinator-plant interactions, landscape proportion of green area and the self-perceived mental health indices of people living close to greenspaces on university campuses and in the host cities; develop a nature-based intervention using findings from the first three objectives.

## Materials and Methods

### Study Location

Five university campuses and their host cities will be surveyed in southwest Nigeria, within the West African Guinea Biodiversity Hotspot, one of the richest biodiversity hotspots in the world with several endemic species [9]. Surveys will be carried out in the following universities and host cities; Obafemi Awolowo University, Ile-Ife; University of Ibadan, Ibadan; Federal University of Technology, Akure; Redeemers University, Ede; Adeyemi Federal University of Education, Ondo. These universities have greenspaces integrated within the academic and administrative areas as well as the staff and student residential areas.

### Study Design

This study is mixed-methods research that is divided into four work packages (WP) based on the outlined objectives.

#### WP 1: Survey of pollinator and plant diversity and landscape proportion of green area as indicators of ecosystem health

In each of the selected five university campuses and host cities, at least two study sites will be identified per university campus or host city. Study sites will be public greenspaces such as playgrounds, parks and gardens. In each sampling location, a site will be defined as a fixed 25 m x 25 m plot where sampling of plants and pollinators and observation of plant-pollinator interactions will be conducted. Fieldwork will be conducted quarterly, to ensure that sampling is conducted in rainy and dry seasons over a one-year period.

During the period of observation, the identity of every insect and flower visited, as well as the number of visits the insect made to the plant within each 25 m x 25 m study site will be noted and recorded. Furthermore, the identity of every plant in each site will be determined, and flower abundance of each plant will be estimated by counting individual flower units. Further, pan traps (bowls of 1 L capacity) of colour white, yellow and blue (a total of twelve bowls per site) will be used to collect diverse insect pollinators in each study site. The bowls will be half-filled with water and few drops of liquid detergent (to break surface tension and enhance insect trapping), and will be deployed in each site for a 24-hour period according to [10]. Insects caught in pan traps will be preserved in 75% ethanol until sorting and identification of insect pollinators. Sampled plants will be identified at the herbarium of Obafemi Awolowo University (OAU) and herbarium collections of other universities while insect pollinators will be identified using identification keys such as [11,12,13] and with reference collection at the AG Leventis Museum of Natural History Obafemi Awolowo University, Ile-Ife and Museum collections of other universities.

The landscape proportion of green area for each study site will be determined with Landsat images using Geographic Information System (ArcGIS Pro Environmental Systems Research Institute, ESRI, California). This will be computed across a gradient of nearness to each study site at 250 m, 500 m, 1000 m, 2000 m and 4000 m radii around each study site.

##### Data Analysis

Pollinator-plant interaction data will be used to construct plant-pollinator networks using the bipartite R package [14]. Quantitative networks can be interpreted using network metrics derived from the estimates of interaction strength and frequency [15]. Plants, pollinators and landscape variables will be analysed using Generalized Linear Mixed Effect Models and Permutation Analysis of Variance (PERMANOVA). All analysis will be conducted in R.

#### WP 2: Survey of mental health indicators

This will involve a cross-sectional survey and a comparative study.

##### Sample Size Determination

The minimum sample size will be determined using the formula for calculating sample size for the comparison of two independent proportions. Based on a WHO Report [16], the prevalence of mental wellbeing in Nigeria was 80% and it is assumed that this study will be able to detect about 15% difference between the prevalence of wellbeing among adults in the university community and adult within the host city. Therefore, the estimated minimum sample size per group will be 256 x 2 = 512. Hence, in the five universities campuses and the respective host cities, we will sample 2560 respondents.

##### Study Population

The respondents will be recruited from the five university campuses and the respective host cities. The respondents that meet the inclusion criteria and give written consent will be randomly recruited until the required number is complete in each study location. We will control for age, sex and level of education between the comparative groups.

Participants will be selected according to the criteria given below:

Inclusion criteria will be as follows:

(i) Participants to be recruited will be individuals aged 18 years and above living within the university and those living in the host community.
(ii) They must have been living for at least 12 months within the study areas.

Exclusion criteria will be as follows:

(i) Individuals aged less than 18 years, those living less than 12 months within the study areas.
(ii) Those who do not give consent

##### Measures

Data will be collected on respondents sociodemographic details, general health and wellbeing, perception of greenspace, multidimensionality of wellbeing using 23-item version of PERMA wellbeing scale [17] and mental health indicators which include depressive symptoms using the Patient Health Questionnaire (PHQ-9) [18]; anxiety symptoms using the Generalized Anxiety Disorder 7-item (GAD-7) [19]; perceived stress using Short Form Perceived Stress Scale (PSS-4) [20]. In addition, suicidal ideation and social connectedness will be measured using Suicidal Ideation Attributes Scale (SIDAS) [21] and Social Connectedness Questionnaire-Revised [22] respectively.

#### WP 3: Analysis of relationship between ecosystem health and mental health

This will involve both qualitative and quantitative assessments. For the qualitative aspect, a non-probabilistic purposive sampling approach will be used.

##### Sample Size Determination

Based on previous studies, in-depth interview may reach a theoretical saturation point after interviewing between 6-8 individuals with average knowledge about the subject matter [23]. From the foregoing, about 6 – 8 participants per study site and a total of 30 – 40 participants will be interviewed.

##### Participants

The respondents will be recruited from the five university campuses and the respective host cities. The respondents that meet the inclusion criteria and give written consent will be recruited. Participants will be selected according to the criteria given in WP 2.

##### Interview guide

The interview guide will explore the social and ecological perceptions of respondents on greenspace. It will further examine interactions of people with specific indicators of ecosystem health in greenspace such as pollinators and plants. Also, we will examine the psychosocial importance of greenspace on respondents self-perceived wellbeing and mental health.

##### In-depth interview

Each respondent will be invited to take part in the project and information about the project will be provided. Informed consent will be obtained, and they will be interviewed using the interview guide. This will be done in a quiet space and privacy will be maintained.

##### Data analysis

The recorded interviews will be transcribed verbatim into Microsoft Word files and uploaded into the NVivo 12 for analysis. Data analysis will be guided by the conventional approach of qualitative content analysis. The quantitative aspect of WP3 will involve scientific analysis using appropriate statistical models to assess the relationship between indicators of ecosystem health and mental health using quantitative data obtained on pollinator and plant diversity in WP 1 and quantitative data obtained on mental health in WP 2.

#### WP 4: Greenspace intervention study

This study will comprise a non-randomized single-arm trial feasibility test and will utilise a mixed methods design utilising qualitative (interviews) methods and quantitative assessments. There will be no control group in this study as the primary focus is on intervention content and feasibility of intervention delivery rather than effectiveness.

##### Sample size

Approximately 60-80 individuals aged 18 years above will be recruited to participate in WP 4 and this is in keeping with other non-randomised feasibility studies. Participants will be recruited from one of the study locations.

##### Greenspace intervention

Green space interventions hold potential benefits for mental and physical health; however, interventions require an integrated approach which harness multidisciplinary skills for a best-fit intervention framework [24]. Based on results obtained in WP 1-3, specific indicators of ecosystem health such as pollinator and plant species which contribute positively to improved mental health will be identified. These will be adapted for the most appropriate intervention/nature-based therapy known as greenspace therapy. A three-to-four round modified Delphi approach will be conducted among a panel of experts with experience in the field of Anthropology, Mental Health, Zoology, Botany, and Public Health. The panel will be presented with the preliminary summaries of WP1, WP2 and WP3. The greenspace intervention will first be independently scored for effectiveness (how clinically effective they think the intervention will be), feasibility (how practicable will it be to implement the intervention in the local setting), and acceptability (the appropriateness of the intervention and it’s fit with the local habits). Each will be scored on a 5-point scale. The experts will later decide on intervention options that can be adaptable to the local environment as greenspace intervention based on brevity, cost and best mode of delivery and total number of sessions. This will be discussed by the panel and the agreement will be by consensus. The developed greenspace intervention will be delivered (the content and session number will be identified during the development phase) to identified individuals. Feasibility and acceptability of the therapy will be assessed using pre-post quantitative evaluation and a post-intervention qualitative examination of participant experiences.

#### Study schedule/timeline

Field surveys and data collection for WP 1 commenced in August 2024. Field survey and participants recruitment for WP 2 and qualitative aspects of WP 3 will start on 20^th^ January 2025 and end on 30^th^ June 2025. Preliminary findings from these WPs will be used for the quantitative aspect of WP 3 and for the modified Delphi panel approach in WP 4. Final data analysis and scientific reporting are expected to be concluded by the mid of 2026 (Table 1).

**Table 1:** Project activity plan and timeline.

| S/N | Description Of Activity | Duration | Year | Jan | Feb. | Mar. | Apr. | May | Jun | Jul | Aug | Sep. | Oct. | Nov. | Dec. |
| --- | --- | --- | --- | --- | --- | --- | --- | --- | --- | --- | --- | --- | --- | --- | --- |
| 1 | Team Meeting for project planning and commencement | One day | 2024 |  |  |  |  |  |  | X |  |  |  |  |  |
| 2 | **Work package 1 Activities** – First round of field work | Two months | 2024 |  | X | X |  |  |  |  |  | X | X |  |  |
| 3 | Sorting, Identification and Digitization of samples | Six weeks | 2024 |  |  |  |  |  |  |  |  |  | X | X |  |
| 4 | Second round of field work | Two months | 2024-2025 | X |  |  |  |  |  |  |  |  |  |  | X |
| 5 | Sorting, Identification and Digitization of samples | Six weeks | 2025 | X | X |  |  |  |  |  |  |  |  |  |  |
| 5 | Third round of field work | Two months | 2025 |  |  | X | X |  |  |  |  |  |  |  |  |
| 6 | Sorting, Identification and Digitization of samples | Six weeks | 2025 |  |  |  | X | X |  |  |  |  |  |  |  |
| 7 | Fourth round of field work | Two months | 2025 |  |  |  |  |  | X | X |  |  |  |  |  |
| 8 | Sorting, Identification and Digitization of samples | Six weeks | 2024-2025 |  |  |  |  |  |  | X | X |  |  |  |  |
| 9 | Data Analysis and reporting | Two months | 2025 |  |  |  |  |  |  |  | X | X |  |  |  |
| 10 | **Work package 2 Activities** – Questionnaire testing and production | Two months | 2024 |  |  |  |  |  |  |  |  |  |  | X | X |
| 11 | Participant recruitment and field survey | Six months | 2025 | X | X | X | X | X | X |  |  |  |  |  |  |
| 12 | Data analysis and reporting | Two months | 2025 |  |  |  |  |  |  | X | X |  |  |  |  |
| 13 | **Work package 3 Activities** –Qualitative survey-Interview questions testing and production | Two months | 2024 |  |  |  |  |  |  |  |  |  |  | X | X |
| 14 | Participant recruitment and field survey | Six months | 2025 | X | X | X | X | X | X |  |  |  |  |  |  |
| 15 | Data analysis and reporting | Two months | 2025 |  |  |  |  |  |  | X | X |  |  |  |  |
| 16 | Quantitative Survey – Analysis of data from WP 1 & WP 2 and reporting | Two months | 2025 |  |  |  |  |  |  |  |  | X | X |  |  |
| 17 | **Work package 4 Activities – Modified Delphi Expert Panel** | Two months | 2025 |  |  |  |  |  |  |  |  |  |  | X | X |
| 18 | Intervention Study | Three months | 2026 | X | X | X |  |  |  |  |  |  |  |  |  |
| 19 | Data Analysis and reporting | Two months | 2026 |  |  |  | X | X |  |  |  |  |  |  |  |
| 20 | Dissemination Workshop | One day | 2026 |  |  |  |  |  | X |  |  |  |  |  |  |

##### Ethics

Ethical clearance was obtained from The Health Research Ethics Committee, Institute of Public Health, Obafemi Awolowo University, Ile-Ife, Nigeria with ethics certificate number: IPH/OAU/12/2413. All study sites were listed in the research proposal submitted for ethical approval which outlined a similar study protocol for all the study sites. According to the ethics guidelines, all study participants will read and sign a written informed consent to confirm they willingly consent to be recruited for the study.

## Discussion

This study aims to provide information on the prospects of academic greenspaces in promoting ecosystem and human health. The potential of these greenspaces in offering nature-based therapy that is adaptable for secondary cities in low– and middle-income countries like Nigeria is being examined. Urban greenspaces can be crucial for maintaining and enhancing both healthy ecosystems and human population in cities [25,26]. Large cities in the global north have a long history, from around the 19th century, when industrialization was accompanied by the establishment of greenspaces in urban areas. In contrast, the expansion of cities in certain developing and tropical countries is more recent and less sustainably designed. Some of these nations frequently have poor urban planning due to unlawful construction and inadequate execution of planned greenspaces, which leaves most urban cities with little to no greenspace [27]. Hence, the goal of this study is to also provide timely scientific information that will help us create awareness programs and workshops about the value of preserving greenspaces in the core and buffer areas of urban ecosystems, aimed at relevant policy makers and particular population groups. This becomes even more imperative in developing nations such as Nigeria with burgeoning population, rapid urbanization [28], few greenspaces [29] and low awareness towards mental health issues [30].

Additionally, the person-environment-health framework—which is based on the Attention Restoration Theory (ART) and Sense of Place Theory (SPT)—is incorporated into this study [31,32,33;34]. It provides a convergent lens for collaborative inquiry into the ways that environments structure and shape human wellbeing [35,36]. It is suggested by the ART that natural settings can facilitate recovery from mental health conditions through subtly fascinating stimuli that are compelling without mental effort [33]. The ability to direct attention to one stimulus at a time (e.g., a task) requires inhibition of competing stimuli and that, over time, this capacity fatigues, resulting in mistakes, failure to focus, or impatience. Another framework for understanding how people interact with urban greenspace is called sense of place, and it proposes that the psychological significance of the place itself cannot be underestimated [31]. Place-related theory and research center on two basic concepts: identification which is the contribution of a location to an individual’s personal identity and attachment which is the emotional connections that people have with a physical place [32] (Fig 1).

**Figure I:**
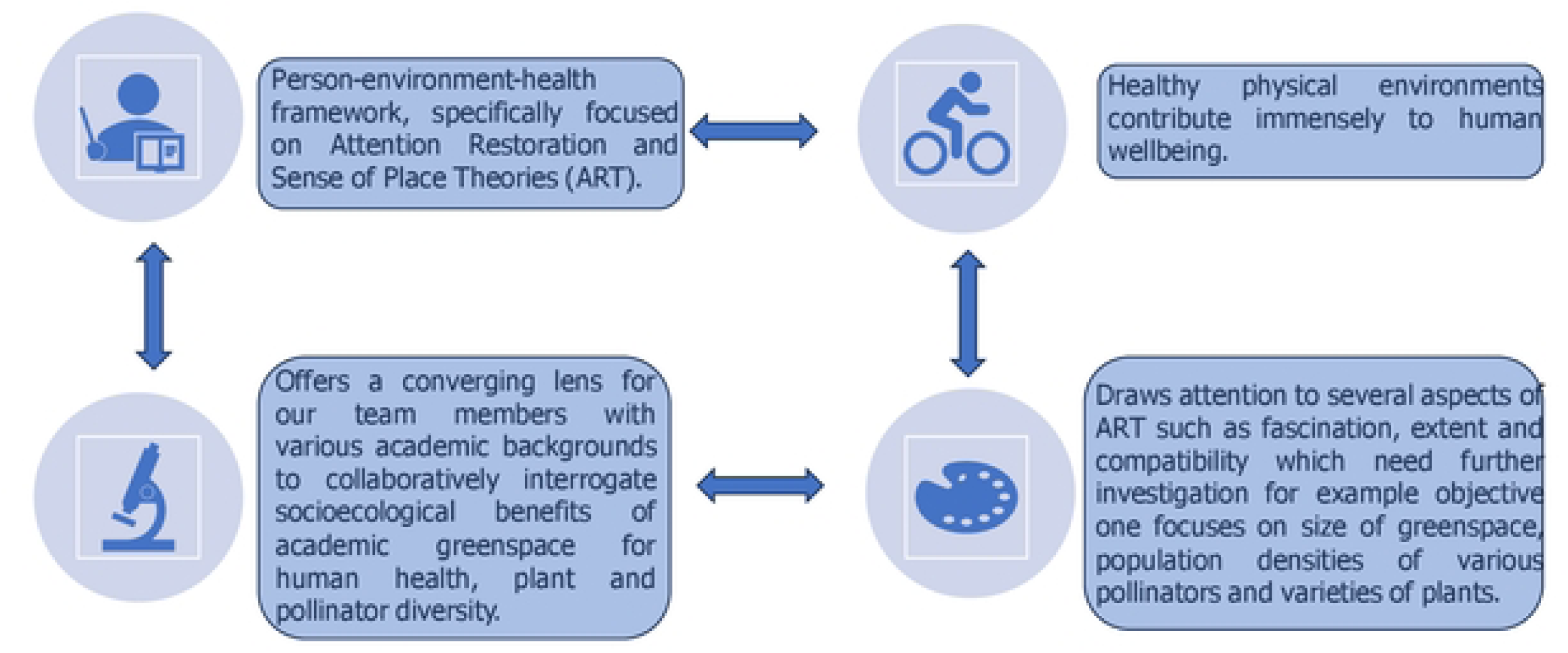
Conceptual framework of the study.

There is increasing evidence that greenspace is often associated with improved human wellbeing [37,38], however, the quality of greenspaces is largely dependent on the availability of ecosystem service providers resident within the urban matrix. Pollinators are critical indicators of ecosystem health through their role in maintaining the diversity of approximately 90% flowering plants globally [39], many of which are important food sources for the growing human population [40,41]. Furthermore, while the contribution of pollinators and flowering plants to critical ecosystem services such as pollination and food security has been well documented [40,41], there is dearth of information on their potential contribution to human health especially in the context of greenspaces in urban areas. Moreover, studies have independently reported the potential benefits of urban greenspaces for pollinators through the provision of food and nesting resources [1,42] and for the mental health of urban populations by reducing emotional and physiological stress through contact with nature and promotion of physical activity and socialization [43,44,45]. In conclusion, this project will explore the potential for inclusive and sustainable nature-based therapy by utilising the socioecological environment that exists in academic greenspaces located in secondary cities. Experiments in nature-based therapy will be carried out with participants of various ages, genders, and socioeconomic backgrounds in order to develop a useful framework that is suitable and beneficial for the use of academic green spaces to support inclusive and sustainable nature-based health benefits for secondary city dwellers. This will make a significant contribution to the promotion and accomplishment of the World Health Organization’s goal of healthy and smart city initiatives [46].

## Limitations

The anticipated limitation of this study is the likely variation in demographics of study participants in the universities compared to the host cities. We plan to address these limitations by ensuring wide spectrum of study participants ranging from students, middle-class workers and non-literate workers living both in the universities and the host cities are recruited during the study.

## Data Availability

No datasets were generated or analysed during the current study. All relevant data from this study will be made available upon study completion.

## Acknowledgements

We acknowledge the gardens and park managers for granting us access to use the sites.

## Notes

### Competing Interest Statement

The authors have declared no competing interest.

### Funding Statement

Yes

### Author Declarations

The Health Research Ethics Committee of the Institute of Public Health at Obafemi Awolowo University, Ile-Ife, Nigeria approved this study. An approval letter from the Committee was issued and the approval number for the study is IPH/OAU/12/2413. The universities Obafemi Awolowo University, Ile-Ife University of Ibadan, Ibadan Federal University of Technology, Akure Redeemers University, Ede and Adeyemi Federal University of Education, Ondo also approved the field research in their universities.

